# Place of Birth and Cognition among Older Americans: Findings from the Harmonized Cognitive Assessment Protocol

**DOI:** 10.1101/2023.10.12.23296954

**Authors:** Zhuoer Lin, Xi Chen

## Abstract

**Objectives:** Growing evidence suggests that place of birth (PoB) and related circumstances may have long-lasting and multiplicative contributions to various later-life outcomes. However, the specific contributions to different domains of cognitive function in late life remain less understood. This study aimed to investigate the extent to which PoB contribute to a wide range of later-life cognitive outcomes.

**Methods:** A nationally representative sample of Americans aged 65 and older (N=3,216) from the Health and Retirement Study (HRS) Harmonized Cognitive Assessment Protocol (HCAP) was utilized. Cognitive outcomes were assessed in HCAP and linked to HRS state-level PoB data to explore the contribution of birthplace to later-life cognitive disparities. Regression-based Shapley decompositions were employed to quantify this contribution.

**Results:** PoB significantly contributed to all assessed cognitive outcomes including memory, executive function, language and fluency, visuospatial function, orientation, global cognitive performance, cognitive impairment and dementia. Geographic disparities in cognitive outcomes were evident, with individuals born in US southern states and foreign-born individuals performing worse than those born in other states. PoB overall accounted for 2.4-13.9% of the total variance in cognition after adjusting for age and sex. This contribution reduced by half when adjusting for a rich set of sociodemographic and health factors over the life course, but PoB still independently explained 2.0-7.1% of the total variance in cognition.

**Discussion:** PoB has lasting contributions to later-life cognitive health, with significant geographic disparities observed. Addressing these disparities requires promoting more equalized place-based policies, resources, and early-life environments to improve health equities over the life course.

## Introduction

Place of birth (PoB hereafter) can contribute profoundly to a host of later-life outcomes. It has been shown to affect a variety of outcomes across the life course from educational attainment, income, to health and well-being (Bifulco et al., 2011; Chetty & Hendren, 2018; Ludwig et al., 2012). As individuals do not get to “choose” where they are born, PoB is often considered as a critical and exogenous source of inequalities and disparities later in life. The socioenvironmental and contextual factors individuals experienced during early life stages, for example, significantly shape human capital development and can have lasting and multiplying influences throughout their lifetimes (Bifulco et al., 2011; Borenstein & Mortimer, 2016; Chetty & Hendren, 2018; Livingston et al., 2020; Ludwig et al., 2012). Understanding the role of PoB in shaping later-life outcomes is crucial for implementing early interventions and effective policymaking.

While the long-term contribution of PoB on health can be substantial, there is limited direct evidence linking birthplace to later-life health outcomes among older adults. Prior studies in the US have indicated that a person’s birthplace is associated with varying levels of mental health, cardiovascular health, chronic diseases, cognition, and mortality rates in adulthood (Fang et al., 2018; Leventhal & Brooks-Gunn, 2003; Rehkopf et al., 2015; Xu et al., 2020). However, most of these studies focus on broad aspects of birthplace, such as rurality (Contador et al., 2015; Lundberg et al., 2009), regions with high disease or mortality risks (Fang et al., 2018; Gilsanz et al., 2017; Glymour et al., 2009), or countries of birth (Fang et al., 2018; Garcia et al., 2019), with only few studies examining more specific differences, such as the state of birth (Brown et al., 2019; Komro et al., 2016; Xu et al., 2020). Importantly, despite the large and persistent disparities in late-life cognitive function in the US, no study to date has explicitly quantified the long-term contribution of the state of birth to cognitive outcomes among older adults. It remains unclear to what extent the state of birth may contribute to late-life disparities in cognition.

State of birth could explain cognitive disparities through several important pathways. First, state policies, resources, and social environments significantly shape early-life exposures, particularly during the prenatal and early childhood stages, which are critical periods for brain and cognitive development (Brown et al., 2019; Komro et al., 2016; Lin & Chen, 2021; Strully et al., 2010; Xu et al., 2020; Zhang et al., 2008, 2016). Differences in state-level health insurance coverage, access to healthcare resources, and income subsidies can directly shape prenatal and postnatal health, nutrition, and cognitive development, leading to varying cognitive performance as individuals age (Brown et al., 2019; Komro et al., 2016; Strully et al., 2010; Xu et al., 2020; Zhang et al., 2008). Second, state-level socioeconomic conditions may create disparate educational opportunities and social environments (e.g., housing, safety), influencing individuals’ ability to build cognitive reserve and pursue cognitive-stimulating occupations that can buffer against cognitive decline in later life (Borenstein & Mortimer, 2016; Lin & Chen, 2021; Zhang et al., 2008, 2016). Education, in particular, has been identified as a crucial factor related to cognition in later life (Livingston et al., 2020), primarily driven by regional differences in education quality and duration (Livingston et al., 2020; Soh et al., 2023). Third, the early-life circumstances shaped by state contexts can contribute significantly to individuals’ health and health behaviors over the life course, which are in turn linked to long-term cognitive health in older age (Alzheimer’s Association, 2023; Borenstein & Mortimer, 2016; Lin & Chen, 2021; Livingston et al., 2020; Zhang et al., 2010). Lastly, structural disadvantages and adversities linked to the state of birth, such as poverty, residential and school segregation, and discrimination, may increase vulnerability to diseases and cognitive disorders among minority groups (Alzheimer’s Association, 2023; N. Jones et al., 2021). Racial and ethnic differences originating from early-life environments may further contribute to later-life differences in cognition (Alzheimer’s Association, 2023; Glymour & Manly, 2008).

To explore the long-term contribution of birth location to cognitive function among Americans aged 65 and older, we linked comprehensive cognitive assessments from the Health and Retirement Study (HRS) Harmonized Cognitive Assessment Protocol (HCAP) with historical geographic data from the HRS. Our analysis revealed substantial geographic variations related to birthplace across a wide range of cognitive outcomes, including memory, executive function, language and fluency, visuospatial function, orientation, global cognitive performance, cognitive impairment, and dementia. We found that older adults born in southern states and foreign countries exhibited poorer cognitive performance than others, and this pattern remained consistent across various cognitive measures.

Using Shapley value decomposition, we demonstrated that PoB accounted for 2.4% to 13.9% of the total variation in cognitive outcomes, after adjusting for age and sex, underscoring its significant contributions. Furthermore, we revealed that PoB independently contributed to 2.0% to 7.1% of the total variation in cognition, even after accounting for a rich set of factors over the life course. This finding suggests that half of the contribution of birthplace is potentially mediated through later-life observable differences in demographics, socioeconomic status (SES), and health factors, with the remaining half being the direct contribution of birthplace. Considering the fact that PoB could independently contribute to 13-21% of the total variation explained by all included factors, which was comparable to the independent contribution of demographic factors including race/ethnicity, marital status and number of children, the contribution of birthplace appears crucial. Overall, our study underscores the lasting contribution of PoB on disparities in cognition later in life, highlighting the importance of public policies in mitigating the disparities related to factors associated with birth states.

## Methods

### Data and Study Participants

The data for this study were obtained from the Health and Retirement Study (HRS), a nationally representative study of American adults aged 50 and older. The HRS has been conducted biannually since 1992, with approximately 19,000 participants interviewed in each wave. In 2016, the HRS developed and conducted the Harmonized Cognitive Assessment Protocol (HCAP) to obtain a comprehensive assessment of cognitive function and dementia risk among older adults (R. N. Jones et al., 2023; Langa et al., 2019; Manly et al., 2022). A total sample of 3,496 older adults aged 65 and older were randomly selected from the 2016 core survey and a comprehensive set of cognitive measures were collected from the respondents and their knowledgeable informants (R. N. Jones et al., 2023; Langa et al., 2019; Manly et al., 2022).

Figure 1 illustrates the sample selection process. After excluding 12 participants without PoB data from the HRS restricted cross-wave geographic data file, 3,484 older adults were included. Subsequently, 223 participants without complete measures of sociodemographic and health factors were further excluded. Finally, a total of 3,261 participants aged 65 and older with complete data on PoB, demographics, SES, and health factors over the life course were included in the analysis.

**Figure 1.**
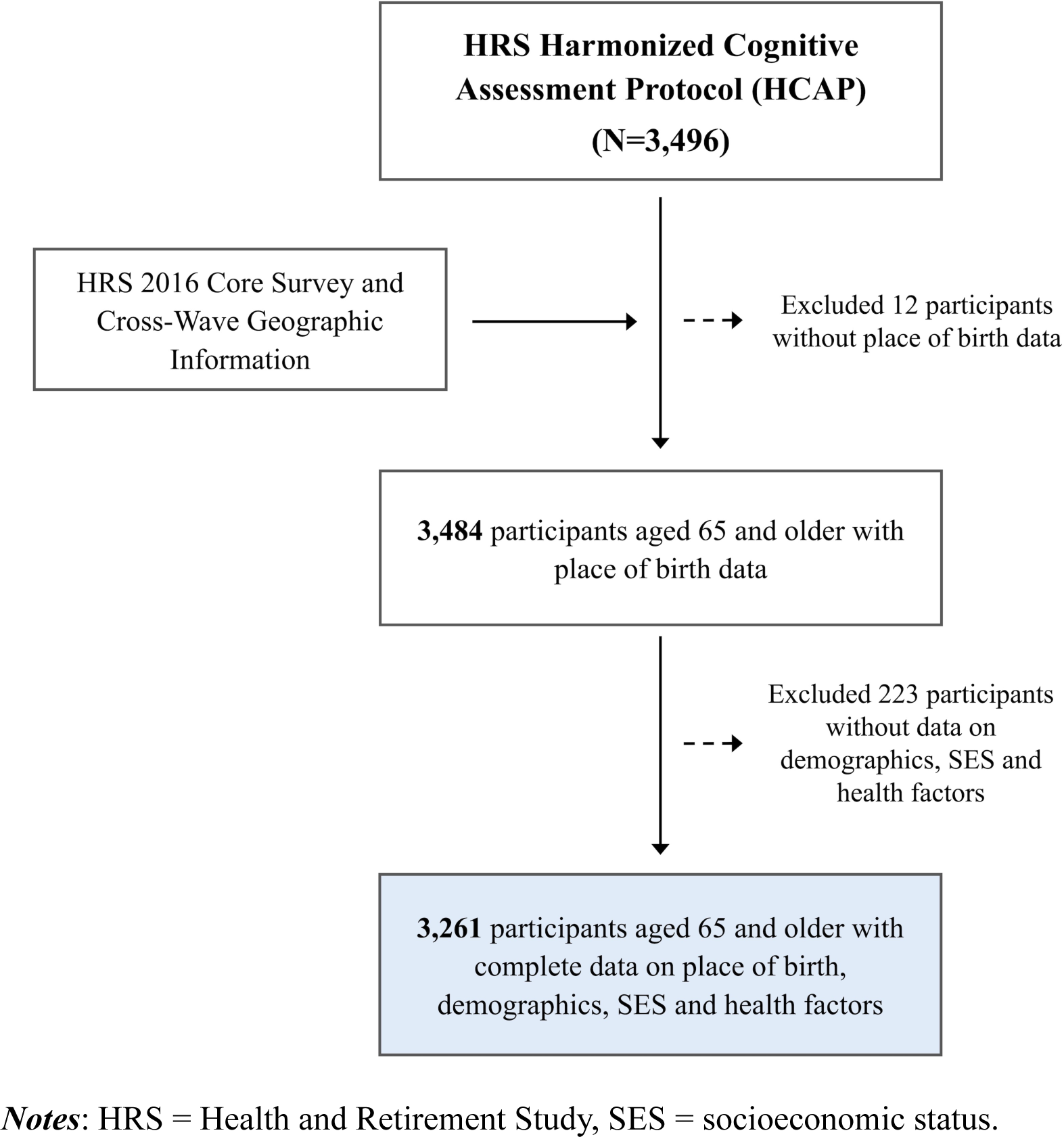
Flow chart of sample selection process

**Table 1.** Characteristics of study participants.

| Characteristics | Descriptive statistics (N=3,261) |
| --- | --- |
| Age, mean (SD) | 75.9 (7.5) |
| Female, n (%) | 1952 (59.9%) |
| **Demographics** |  |
| Race/Ethnicity |  |
| Non-Hispanic White, n (%) | 2343 (71.8%) |
| Non-Hispanic Black, n (%) | 506 (15.5%) |
| Hispanic, n (%) | 340 (10.4%) |
| Other, n (%) | 72 (2.2%) |
| Married/Partnered, n (%) | 1862 (57.1%) |
| Number of Children, mean (SD) | 3.2 (2.0) |
| **Socioeconomic Status (SES)** |  |
| Education in years, mean (SD) | 12.8 (3.1) |
| Wealth level |  |
| Lowest, n (%) | 582 (17.8%) |
| Lower-middle, n (%) | 750 (23.0%) |
| Upper-middle, n (%) | 891 (27.3%) |
| Highest, n (%) | 1038 (31.8%) |
| Working for pay, n (%) | 586 (18.0%) |
| Medicare enrollment, n (%) | 3102 (95.1%) |
| Medicaid enrollment, n (%) | 332 (10.2%) |
| Military health plan enrollment, n (%) | 253 (7.8%) |
| Private health insurance coverage, n (%) | 1291 (39.6%) |
| Employer-based health insurance coverage, n (%) | 707 (21.7%) |
| **Health** |  |
| Hypertension, n (%) | 2295 (70.4%) |
| Diabetes, n (%) | 973 (29.8%) |
| Cancer, n (%) | 653 (20.0%) |
| Lung diseases, n (%) | 443 (13.6%) |
| Heart diseases, n (%) | 1116 (34.2%) |
| Stroke, n (%) | 406 (12.5%) |
| Psychiatric disorders, n (%) | 642 (19.7%) |
| Arthritis, n (%) | 2339 (71.7%) |
| ADL limitations, n (%) | 746 (22.9%) |
| IADL limitations, n (%) | 680 (20.9%) |
| Currently smoking, n (%) | 247 (7.6%) |
| Ever smoking, n (%) | 1797 (55.1%) |
| **Place of Birth (Census Region)** |  |
| New England | 125 (3.8%) |
| Middle Atlantic | 445 (13.7%) |
| East North Central | 581 (17.8%) |
| West North Central | 341 (10.5%) |
| South Atlantic | 488 (15.0%) |
| East South Central | 295 (9.1%) |
| West South Central | 321 (9.8%) |
| Mountain | 98 (3.0%) |
| Pacific | 187 (5.7%) |
| Foreign born | 377 (11.6%) |
| **Cognition** |  |
| Memory, mean (SD) | 50.2 (9.9) |
| Executive function, mean (SD) | 50.2 (9.9) |
| Language and fluency, mean (SD) | 50.2 (10.0) |
| Visuospatial, mean (SD) | 50.2 (10.0) |
| Orientation, mean (SD) | 9.3 (1.3) |
| Global cognitive performance, mean (SD) | 50.3 (9.9) |
| Cognitive impairment, n (%) | 1087 (33.3%) |
| Dementia, n (%) | 343 (10.5%) |

**Notes:** ADL = Activities of Daily Living; IADL = Instrumental Activities of Daily Living. The primary geographic unit for analyzing the impact of place of birth was the state of birth. However, due to restrictions on disclosing summary statistics at the state level, region-level birth location data extracted from the HRS public data file to obtain summary statistics at the U.S. census division level and to map the regional variation in cognitive outcomes. The sample was divided into ten birthplace regions, which included the nine U.S. Census Divisions and foreign-born individuals. The cognitive scores of each domain, except for orientation, were standardized to a mean of 50 and a standard deviation of 10 during the initial construction of domain-specific estimates, and unadjusted versions of the estimates were used in this study.

### Cognitive Outcomes

Cognitive outcomes were assessed using the HRS/HCAP summary cognitive and functional measures data file, which contained processed HRS/HCAP data with cognitive domain-level estimates and dementia and mild cognitive impairment (MCI) classifications (R. N. Jones et al., 2023; Langa et al., 2019; Manly et al., 2022). Domain-level score estimates were derived through factor analyses based on carefully selected cognitive performance items collected in HCAP (R. N. Jones et al., 2023). Dementia and MCI classifications were developed using a dementia algorithm based on HCAP and HRS data (Manly et al., 2022).

For this study, the domain-specific score estimates of five key cognitive domains - memory, executive function, language and fluency, visuospatial function, and orientation - were used as cognitive outcomes, along with the score estimates of global cognitive performance. These scores, except for orientation, were standardized to a mean of 50 and a standard deviation of 10 during their initial construction, and unadjusted versions of the estimates were used in the analysis (R. N. Jones et al., 2023). Additionally, two dichotomous cognitive outcomes were constructed to indicate whether individuals had cognitive impairment (including both MCI and dementia, 0/1) and dementia (0/1) based on an established algorithm developed using HCAP (R. N. Jones et al., 2023; Manly et al., 2022).

### Place of Birth

State-level PoB data were extracted from the HRS restricted cross-wave geographic information data file. Respondents were asked about their PoB during their baseline interview, and those born in the U.S. were further asked which state they were born in. The sample included respondents born in 50 U.S. states, 2 U.S. territories (Washington, D.C., and Puerto Rico), U.S. born without state information, and foreign-born individuals. Foreign-born individuals were classified as such in this study.

The primary geographic unit for analyzing the contribution of PoB was the state of birth. However, due to restrictions on disclosing summary statistics at the state level, region-level birth location data were also extracted from the HRS public data file to obtain summary statistics at the U.S. census division level and to map the regional variation in cognitive outcomes. The sample was divided into ten birthplace regions, which included the nine U.S. Census Divisions and foreign-born individuals. All analyses adhered to the HRS disclosure guidelines, and results generated using HRS restricted data were reviewed and approved by HRS.

### Statistical Analyses

For each cognitive outcome, the Kruskal-Wallis test was used to determine differences in outcomes across PoB after adjusting for age and sex. A significant result would indicate considerable disparities in cognitive performance distributions across different PoB (Kruskal & Wallis, 1952).

To quantify the proportion of variance in outcomes attributed to PoB, we utilized Shapley value decomposition. This method decomposes the explained variance (R2) of a regression model into clusters of explanatory variables that belong to the same domain (Cao et al., 2022; Liu et al., 2019; Shorrocks, 2013; Yan et al., 2020). The regression model used in the analysis includes PoB as the main independent variable, with age, sex, and a comprehensive set of life course factors serving as explanatory variables. The three domains of the later-life factors included demographics, SES and health, which might be partially explained by PoB and subsequently contribute to later-life cognitive outcomes (Alzheimer’s Association, 2023; Alzheimer’s Disease International, 2018; Livingston et al., 2020).

The decomposition analyses were conducted stepwise, adding the three domains of life course factors one by one to examine potential pathways through which PoB may affect late-life cognitive outcomes. In Model A (the most parsimonious setting), we only adjusted for age and sex. In Model B, we further adjusted for demographics, including race/ethnicity, marital status and the number of children. In Model C, we additionally adjusted for SES, including education (measured in years), wealth level (categorized into quartiles), working status, Medicare enrollment, Medicaid enrollment, military health plan enrollment, private health insurance coverage, and employer-based insurance coverage. Lastly in Model D (the most comprehensive setting), we additionally adjusted for health factors, including hypertension, diabetes, cancer, lung diseases, heart diseases, stroke, psychiatric disorders, arthritis, functional limitations for basic or instrumental activities of daily living (ADL/IADL), and smoking behaviors (Alzheimer’s Association, 2023; Alzheimer’s Disease International, 2018; Livingston et al., 2020).

Linear models were used for continuous outcomes (e.g., memory score), and logit models were used for binary outcomes (e.g., dementia). Joint significance tests were conducted to assess the contribution of PoB on each cognitive outcome, and then the decomposition analyses were performed to quantify the contribution of PoB to each cognitive outcome and the contribution of each domain of the life course factors. All statistical analyses were performed using Stata 17.0, and significance was determined at the 5% level (Cao et al., 2022; Liu et al., 2019; Shorrocks, 2013; Yan et al., 2020).

## Results

### Sample Characteristics

Figure 1 presents the characteristics of the sample. Among the 3,216 participants, the mean age was 75.9 years (SD=7.5), and 1,952 (59.9%) were female. The majority of participants were non-Hispanic White (71.8%), and 57.1% were married or had a partner. The mean years of education was 12.8 years (SD=3.1), and 3,102 (95.1%) were covered by Medicare. The most prevalent comorbidities were hypertension (70.4%) and arthritis (71.7%), followed by heart diseases (34.2%) and diabetes (29.8%). Additionally, 55.1% of participants had ever smoked cigarettes in their life, while 7.6% were current smokers. Regarding PoB (census region), 377 (11.6%) of participants were born outside the US (i.e., foreign born). As for cognitive status, 1,087 (33.3%) of participants were identified as having cognitive impairment (including MCI and dementia), and 343 (10.5%) were identified as having dementia.

### Geographic Variation in Cognitive Outcomes by Place of Birth

Figure 2 illustrates the geographic variations in cognitive outcomes by PoB after adjusting for age and sex. Each map shows the average levels of cognitive outcomes at the census division level, with deeper color indicating worse cognitive performance. The *P*-values of Kruskal-Wallis tests are provided at the bottom of each graph, indicating the significance of differences in cognitive outcomes at the state level. Large geographic variations in cognitive outcomes were observed by PoB, and these patterns were consistent across various cognitive domains and outcomes. Specifically, participants born in West South Central, East South Central, and South Atlantic regions, as well as foreign-born individuals, tended to exhibit poorer cognitive outcomes compared to those born in other census regions and divisions. Conversely, those born in the East North Central, Middle Atlantic, and New England regions demonstrated better cognitive performance compared to other birth locations. The Kruskal-Wallis test indicated that the differences in cognitive outcomes across PoB (measured at the state level) were statistically significant for all five cognitive domains and global cognitive performance (*P* <0.001). Additionally, significant differences across PoB were found in the prevalence of cognitive impairment (*P*<0.001) and dementia (*P*<0.001).

**Figure 2.**
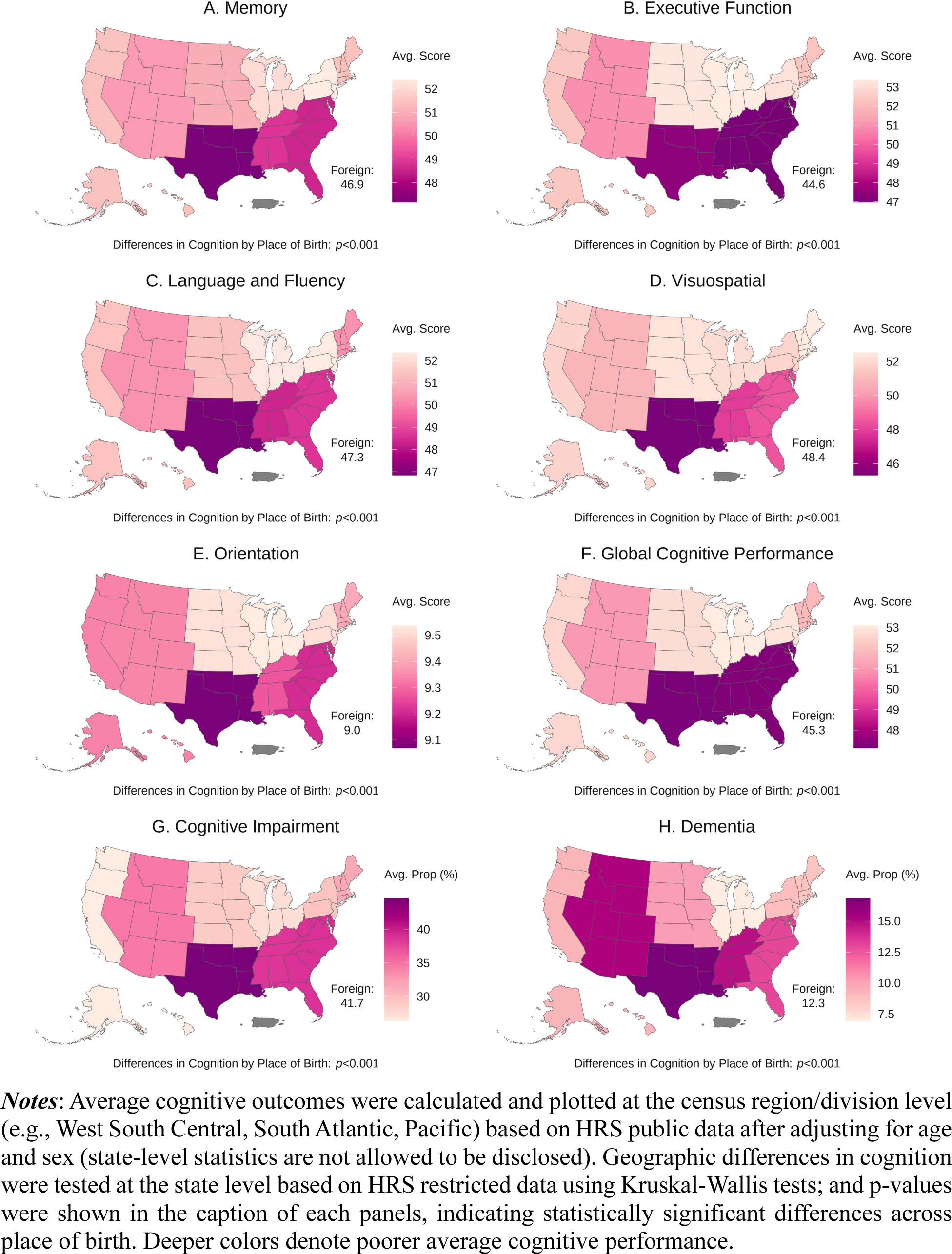
Geographic variation in cognitive outcomes by place of birth

### Contribution of Place of Birth to Later-life Cognitive Outcomes

Figure 3 presents the influence of PoB and the estimates of Shapley decomposition analyses, which quantify the contribution of PoB to the total variance of each cognitive outcome (also see Supplementary Table S1 for the estimates). Panel A shows the contribution of PoB from Model A to Model D. In Model A, which only adjusted for age and sex, PoB accounted for a considerable amount of the total variance in memory (4.9%; significance of joint test: *P*<0.001), executive function (13.9%; *P* <0.001), language and fluency (5.8%; *P* <0.001), visuospatial function (8.0%; *P*<0.001), orientation (3.3%; *P*<0.001), and global cognitive performance (11.2%; *P*<0.001). It also explained 2.4% of the variance in cognitive impairment prevalence (*P*<0.001) and 3.1% of the variance in dementia prevalence (*P* =0.008). As additional life course sociodemographic and health factors were controlled from Model B to D, the contribution of PoB gradually declined, but even in the most comprehensive setting (Model D), it still explained a significant proportion of variance in memory (2.6%; 95% CI, 1.7-3.6%; *P*<0.001 for joint test), executive function (7.1%; 95% CI, 5.8-8.4%; *P*<0.001), language and fluency (3.6%; 95% CI, 2.4-4.8%;*P*<0.001), visuospatial function (5.2%; 95% CI, 3.8-6.6%; *P*<0.001), orientation (2.0%; 95% CI, 0.8-3.3%; *P* <0.001), global cognitive performance (5.7%; 95% CI, 4.4-6.9%; *P* <0.001), cognitive impairment (2.0%; 95% CI, 1.2-2.9%; *P*<0.001), and dementia (2.8%; 95% CI, 1.3-4.2%; *P*=0.039).

**Figure 3.**
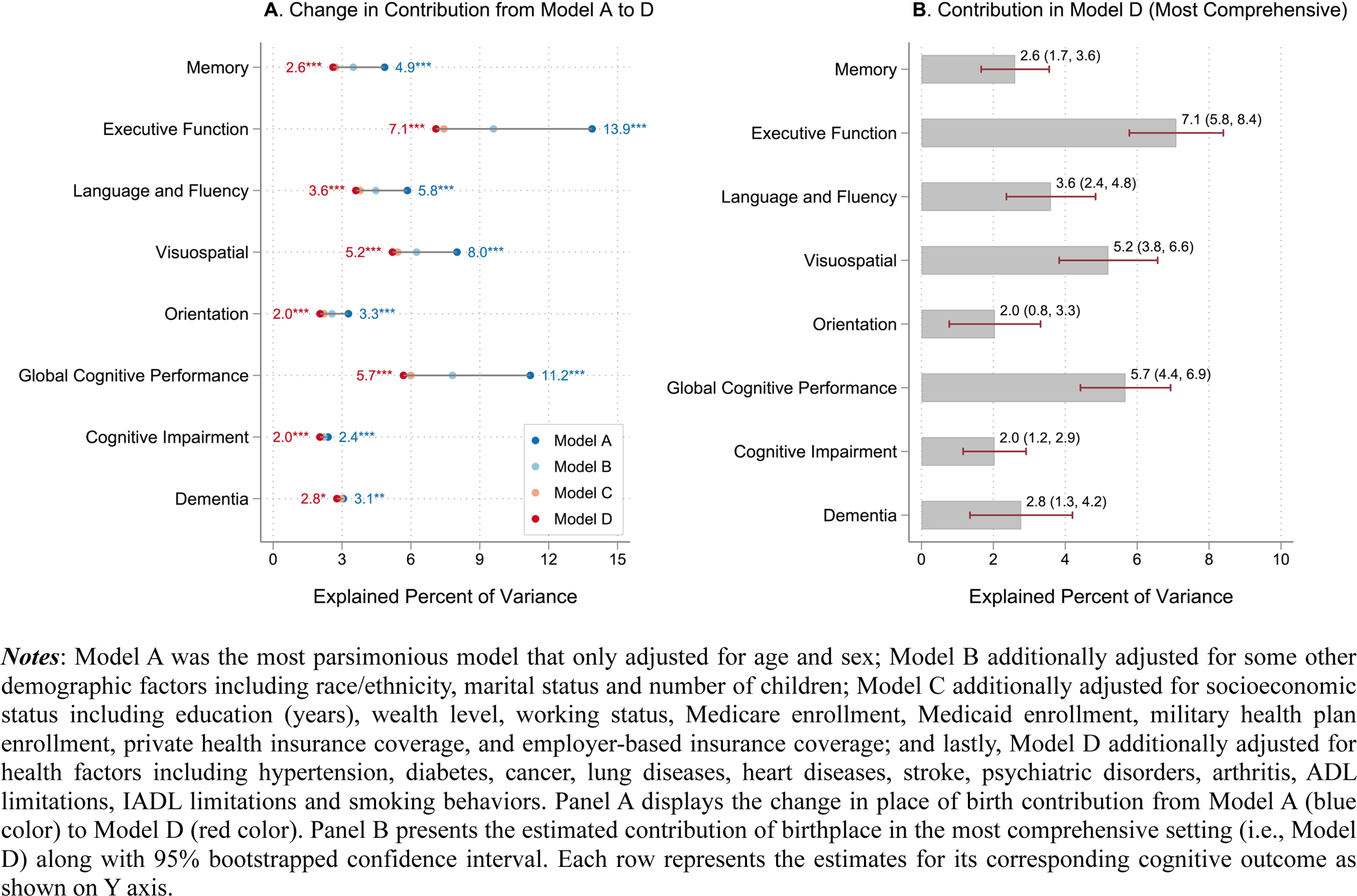
Contribution of place of birth to late-life cognitive outcomes with different model specifications

Figure 4 compares the independent contribution of PoB with those of other life course factors estimated in Model D (also see Supplementary Table S2 for the estimates). The independent contributions of PoB to cognitive outcomes were relatively larger than those of demographic factors but lower than the contributions of SES and health factors. The relative contribution of PoB, expressed in parentheses, ranged from 13% to 21% of the total variance explained by the rich set of factors over the life course, highlighting its clinical significance in contributing to late-life cognitive outcomes.

**Figure 4.**
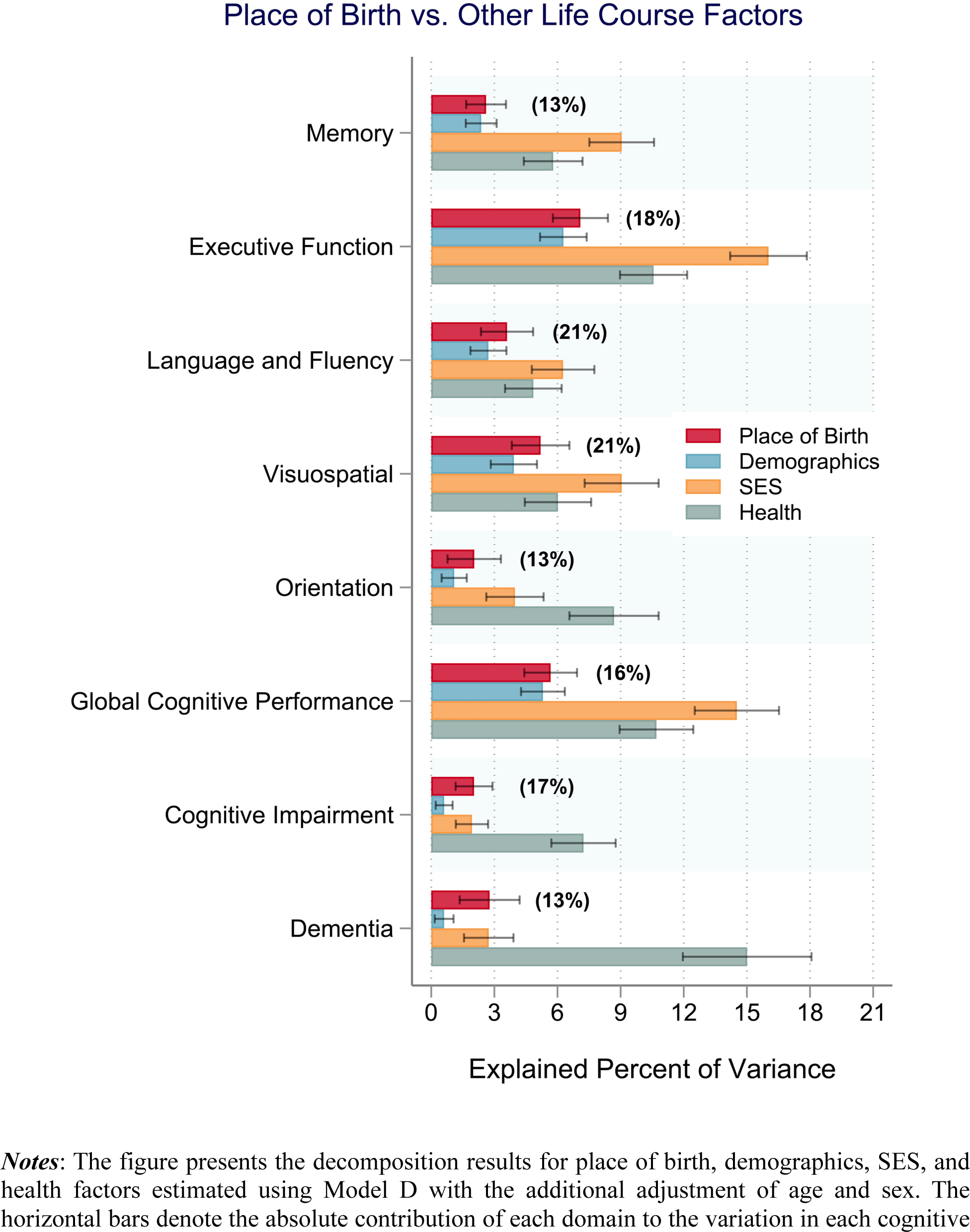
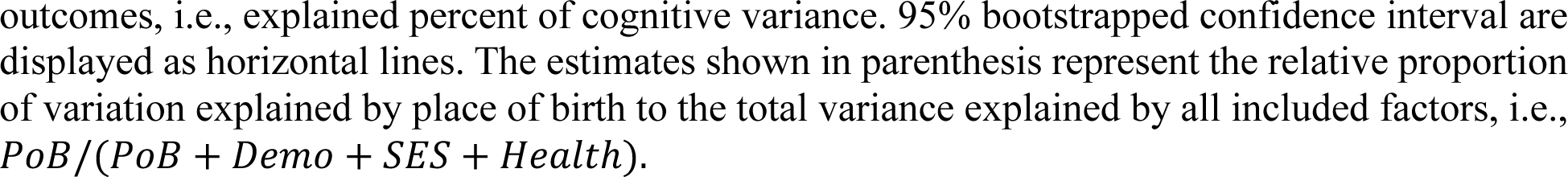
Contribution of place of birth to cognitive outcomes as compared to other life course factors

## Discussion

PoB can have significant and enduring contributions to individuals’ health and well-being throughout their lives. In this study, we provided first evidence on the long-term contribution of state of birth to a range of later-life cognitive outcomes. We found that PoB, measured at the state level, significantly explained cognition, accounting for 2.4-13.9% of the total variance after adjusting for age and sex. We showed that half of these effects were potentially mediated through various life course sociodemographic and health factors, with another half being the independent contribution of birthplaces, highlighting the importance of considering these underlying pathways.

Our findings revealed marked geographic variations in cognitive outcomes across states of birth. People born in certain southern states (in West South Central, East South Central, and South Atlantic) exhibited worse cognitive outcomes compared to those born in other states. This aligns with previous research linking region of birth to cognitive health, such as the higher risk for dementia in the southern Stroke Belt states (Gilsanz et al., 2017; Glymour et al., 2011; Topping et al., 2021). In our study, a comprehensive set of cognitive domains and outcomes were examined in the same context and consistent patterns of disparities were found, which further demonstrates the strength and robustness of the finding. Moreover, individuals born in foreign countries showed poorer cognitive performance than those born in the US, which is consistent with previous evidence (Kovaleva et al., 2021).

We showed that PoB contributed significantly to later-life disparities in cognition. Prior literature suggests that PoB may influence various aspects of individuals’ health and well-being over their life course, including education, income, subjective well-being (Bifulco et al., 2011; Chetty & Hendren, 2018; Ludwig et al., 2012), and various diseases and conditions (Fang et al., 1996, 2018; Gilsanz et al., 2017; Glymour et al., 2009; Lundberg et al., 2009; Patton et al., 2011; Rehkopf et al., 2015; Shiue, 2014; Xu et al., 2020). In our study, accounting for these factors attenuated about half of the contribution of PoB to cognition, emphasizing the importance of addressing these underlying pathways through improved policies affecting education and socioeconomic factors. However, even after accounting for these factors, PoB still independently contributed to later-life cognition. For instance, early-life circumstances shaped by state policies and social environments may contribute directly to brain development and cognitive reserve, leading to long-lasting disparities in later life. Policymakers should consider targeted interventions such as child subsidies and insurance to reduce potential long-term inequities (Brown et al., 2019; Komro et al., 2016; Lin & Chen, 2021; Strully et al., 2010; Zhang et al., 2008, 2016).

Furthermore, while PoB contributed significantly to overall cognitive outcomes, there was some heterogeneity across different cognitive domains. PoB contributed relatively more to domains like executive function and visuospatial abilities but less to domains like memory and orientation. These findings highlight the importance of collecting comprehensive measures of cognitive function, as focusing solely on global cognitive function or specific classification criteria may mask important differences across cognitive domains and underestimate the contribution of certain risk factors on cognitive health (R. N. Jones et al., 2023; Langa et al., 2019; Manly et al., 2022).

Despite these findings, there are some limitations. The sample size based on HCAP data may limit the statistical power for estimating place effects at the state or lower levels. Future studies with larger cohorts could further validate these results. Additionally, due to sample size constraints, we were unable to conduct subsample analyses or further investigate the contribution of migration during early childhood compared to prolonged residence in a specific location. Future studies using residential history data with measured durations could shed more light on these mechanisms. Lastly, recall bias may potentially affect the accuracy of the PoB measure. However, as PoB data were collected during the initial survey when respondents were at younger ages, this concern is somewhat mitigated.

In conclusion, our findings demonstrate the long-lasting and enduring contributions of PoB on later-life cognitive health. Addressing state-level policies, resources, and early-life environments is crucial to improving health equities over the life course. Policymakers should prioritize efforts to equalize opportunities and resources across different states to promote better cognitive health outcomes for all individuals.

## Data Availability

All publicly available data used in the present study are available upon reasonable request to the authors.

## Funding

This work was supported by the Career Development Award [K01AG053408] and a major research grant [R01AG077529] from the National Institute on Aging; Claude D. Pepper Older Americans Independence Center at Yale School of Medicine, funded by the National Institute on Aging [P30AG021342]; and Yale Alzheimer’s Disease Research Center [P30AG066508]; James Tobin Research Fund at Yale Economics Department; and Yale Macmillan Center Faculty Research Award.

## Conflict of interest

We have no conflict of interest to declare.

## Author contributions

Z.L planed the study, performed the data analysis, and wrote the paper. X.C planned the study, supervised the data analysis, and revised the paper.

---

**Supplementary Table S1.**
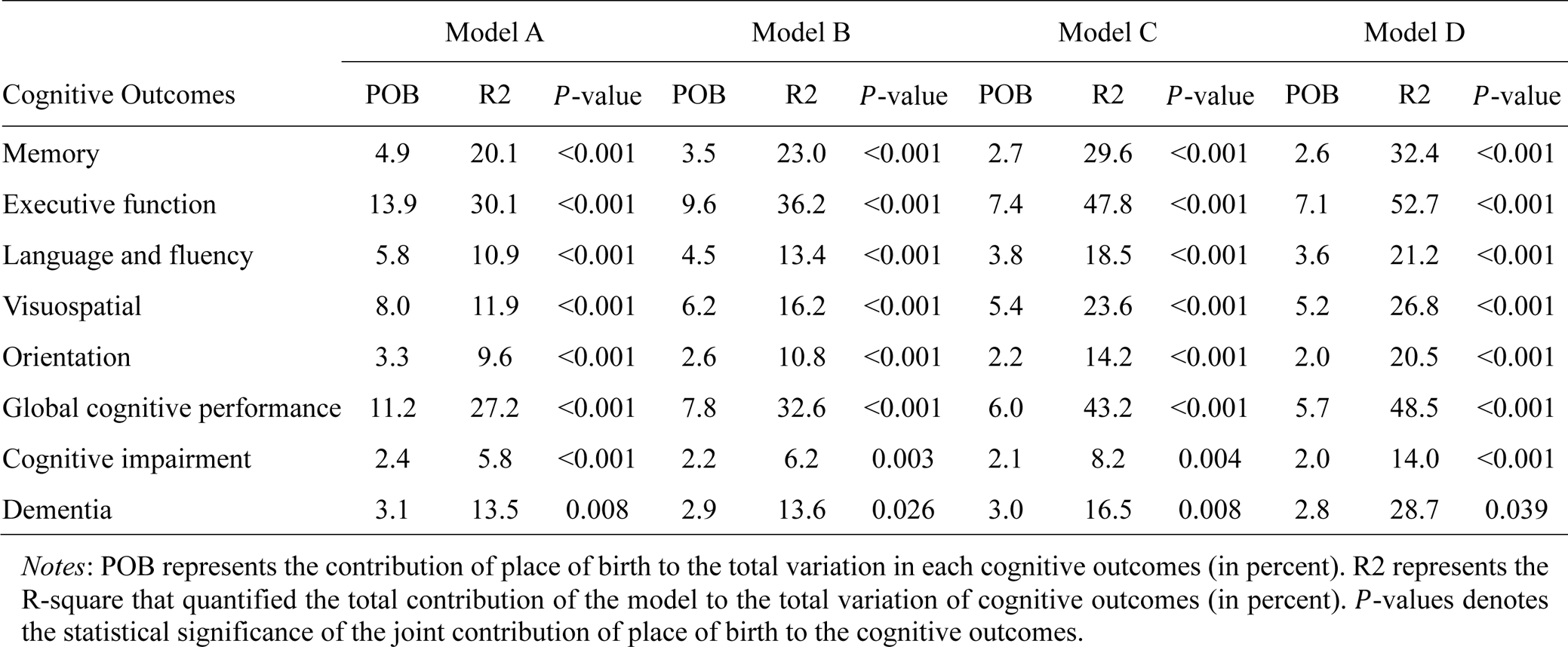
Contribution of Place of Birth to Cognitive Outcomes by Model Specifications.

| Cognitive Outcomes | Model A |  |  | Model B |  |  | Model C |  |  | Model D |  |  |
| --- | --- | --- | --- | --- | --- | --- | --- | --- | --- | --- | --- | --- |
|  | POB | R2 | *P*-value | POB | R2 | *P*-value | POB | R2 | *P*-value | POB | R2 | *P*-value |
| Memory | 4.9 | 20.1 | <0.001 | 3.5 | 23.0 | <0.001 | 2.7 | 29.6 | <0.001 | 2.6 | 32.4 | <0.001 |
| Executive function | 13.9 | 30.1 | <0.001 | 9.6 | 36.2 | <0.001 | 7.4 | 47.8 | <0.001 | 7.1 | 52.7 | <0.001 |
| Language and fluency | 5.8 | 10.9 | <0.001 | 4.5 | 13.4 | <0.001 | 3.8 | 18.5 | <0.001 | 3.6 | 21.2 | <0.001 |
| Visuospatial | 8.0 | 11.9 | <0.001 | 6.2 | 16.2 | <0.001 | 5.4 | 23.6 | <0.001 | 5.2 | 26.8 | <0.001 |
| Orientation | 3.3 | 9.6 | <0.001 | 2.6 | 10.8 | <0.001 | 2.2 | 14.2 | <0.001 | 2.0 | 20.5 | <0.001 |
| Global cognitive performance | 11.2 | 27.2 | <0.001 | 7.8 | 32.6 | <0.001 | 6.0 | 43.2 | <0.001 | 5.7 | 48.5 | <0.001 |
| Cognitive impairment | 2.4 | 5.8 | <0.001 | 2.2 | 6.2 | 0.003 | 2.1 | 8.2 | 0.004 | 2.0 | 14.0 | <0.001 |
| Dementia | 3.1 | 13.5 | 0.008 | 2.9 | 13.6 | 0.026 | 3.0 | 16.5 | 0.008 | 2.8 | 28.7 | 0.039 |

*Notes:* POB represents the contribution of place of birth to the total variation in each cognitive outcomes (in percent). R2 represents the R-square that quantified the total contribution of the model to the total variation of cognitive outcomes (in percent). *P*-values denotes the statistical significance of the joint contribution of place of birth to the cognitive outcomes.

**Supplementary Table S2.** Absolute and Relative Contribution of Place of Birth and Life Course Factors to Cognitive Outcomes in Model D (the Most Comprehensive Model)

| Cognitive Outcomes | Domain | Absolute Contribution | Total Contribution | Relative Contribution |
| --- | --- | --- | --- | --- |
| Memory | Place | 2.6 (1.7, 3.6) | 19.8 | 13.1 |
|  | Demographics | 2.4 (1.6, 3.1) |  | 12.0 |
|  | SES | 9.1 (7.5, 10.6) |  | 45.6 |
|  | Health | 5.8 (4.4, 7.2) |  | 29.2 |
| Executive Function | Place | 7.1 (5.8, 8.4) | 40.0 | 17.7 |
|  | Demographics | 6.3 (5.2, 7.4) |  | 15.7 |
|  | SES | 16.0 (14.2, 17.8) |  | 40.1 |
|  | Health | 10.6 (9.0, 12.2) |  | 26.4 |
| Language and Fluency | Place | 3.6 (2.4, 4.8) | 17.4 | 20.7 |
|  | Demographics | 2.7 (1.9, 3.6) |  | 15.6 |
|  | SES | 6.3 (4.8, 7.8) |  | 35.9 |
|  | Health | 4.9 (3.5, 6.2) |  | 27.8 |
| Visuospatial | Place | 5.2 (3.8, 6.6) | 24.2 | 21.5 |
|  | Demographics | 3.9 (2.8, 5.0) |  | 16.2 |
|  | SES | 9.0 (7.3, 10.8) |  | 37.4 |
|  | Health | 6.0 (4.4, 7.6) |  | 24.9 |
| Orientation | Place | 2.0 (0.77, 3.3) | 15.8 | 12.9 |
|  | Demographics | 1.1 (0.49, 1.7) |  | 6.9 |
|  | SES | 4.0 (2.6, 5.3) |  | 25.2 |
|  | Health | 8.7 (6.6, 10.8) |  | 55.0 |
| Global Cognitive Performance | Place | 5.7 (4.4, 6.9) | 36.2 | 15.7 |
|  | Demographics | 5.3 (4.3, 6.3) |  | 14.7 |
|  | SES | 14.5 (12.5, 16.5) |  | 40.1 |
|  | Health | 10.7 (9.0, 12.5) |  | 29.6 |
| Cognitive Impairment | Place | 2.0 (1.2, 2.9) | 11.8 | 17.2 |
|  | Demographics | 0.61 (0.21, 1.0) |  | 5.2 |
|  | SES | 1.9 (1.2, 2.7) |  | 16.4 |
|  | Health | 7.2 (5.7, 8.8) |  | 61.2 |
| Demented | Place | 2.8 (1.3, 4.2) | 21.1 | 13.1 |
|  | Demographics | 0.61 (0.17, 1.1) |  | 2.9 |
|  | SES | 2.7 (1.6, 3.9) |  | 12.9 |
|  | Health | 15.0 (12.0, 18.1) |  | 71.0 |

**Notes:** The table presents the decomposition results for place of birth, demographics, socioeconomic status (SES), and health factors estimated using Model D with the additional adjustment of age and sex (as visualized in Figure 4). Absolute contribution denotes the contribution of each domain to the variation in each cognitive outcomes, i.e., explained percent of cognitive variance. 95% bootstrapped confidence interval are shown in parentheses. Total contribution represents the joint contribution of the four domains to the variation in cognitive outcomes, i.e., $PoB + Demo + SES + Health$ . Relative contribution represents the relative proportion of variation explained by place of birth to the total variance explained by all included factors, i.e., $PoB / (PoB + Demo + SES + Health)$ .

## Notes

### Competing Interest Statement

The authors have declared no competing interest.

